# SARS-CoV-2 Exposures of Healthcare Workers from Primary Care, Long-Term Care Facilities and Hospitals: A Nationwide Matched Case-Control Study

**DOI:** 10.1101/2022.02.26.22271545

**Authors:** Martin Belan, Tiffany Charmet, Laura Schaeffer, Sarah Tubiana, Xavier Duval, Jean-Christophe Lucet, Arnaud Fontanet, Gabriel Birgand, Solen Kernéis

## Abstract

**Objectives:** Healthcare workers (HCWs) are at higher risk of contracting coronavirus disease-19 (COVID-19) than the general population. This study assessed the roles of various exposures and personal protective equipment (PPE) use on that risk for HCWs working in primary care, long-term-care facilities (LTCFs) or hospitals.

**Methods:** We conducted a matched case-control (1:1) study (10 April–9 July 2021). Cases (HCWs with confirmed COVID-19) and controls (HCWs without any COVID-19-positive test or symptoms) recruited by email were invited to complete an online questionnaire on their exposures and PPE use. Questions covered the 10 days preceding symptom onset for cases (or testing if asymptomatic) or inclusion for controls.

**Results:** A total of 4152 matched cases and controls were included. The multivariable conditional logistic regression analysis retained exposure to an infected person outside work (adjusted odds ratio, 19.9 [95% confidence intervaI, 12.4–31.9]), an infected colleague (2.26 [1.53–3.33]) or COVID-19 patients (2.37 [1.66–3.40]), as independent predictors of COVID-19 in HCWs, while partial or complete immunization was protective. Eye protection (0.57 [0.37–0.87]) and wearing a gown (0.58 [0.34–0.97]) during COVID-19 patient care were protective, while wearing an apron slightly increased the risk of infection (1.47 [1.00–2.18]). N95-respirator protection was comparable to that of surgical masks. Results were consistent across healthcare-facility categories.

**Conclusions:** HCWs were more likely to get COVID-19 in their personal sphere than during occupational activities. Our results suggest that eye protection for HCWs during patient care should be actively promoted.

## INTRODUCTION

Protecting healthcare workers (HCWs) from coronavirus disease-19 (COVID-19) is critical to ensure their own safety, and maintain continuity and quality of care. HCWs are estimated to have a 1.6- to 3.4-fold higher risk of infection compared to the general population [1, 2]. High on-site involvement during the acute phases of the pandemic and lockdown periods, direct interactions with patients and lack of access to personal protective equipment (PPE) likely contributed to higher exposure. The World Health Organization estimated that between 80 000 and 180 000 HCWs could have died from COVID-19 between January 2020 and May 2021 [3]. In France, from March 2020 to September 2021, 87 647 (9%) laboratory-confirmed infections and 19 attributable deaths were reported among 935 732 HCWs from healthcare facilities [4]. The more recent emergence of highly transmissible variants further affected the healthcare-workforce capacity.

As in the general population, younger male HCWs with comorbidities, in contact with an infected household member or who participated in gathering events are at higher risk for contracting COVID-19 [5–8]. Specific occupational exposures were identified: regular patient-facing activities and contacts with infected colleagues [2, 5, 6, 9, 10]. PPE-conferred protection was mainly studied for Influenza, Severe Acute Respiratory Syndrome (SARS) and Middle East Respiratory Syndrome (MERS), but evidence is controversial for COVID-19 [11–13]. Moreover, very few data have been published on HCWs in long-term care facilities (LTCFs) and primary care [14, 15], despite their intense involvement in the pandemic response. Most studies focused on the hospital setting, although exposures, organization of care and access to infection prevention and control (IPC) expertise vary greatly across facilities.

This study aimed to identify occupational and non-occupational exposures, and PPE practices associated with COVID-19 risk for HCWs working in primary care, LTCFs or hospitals.

## METHODS

### Study Design and Participants

We conducted a matched case-control study from an ongoing national survey (ComCor) led by Institut Pasteur (Paris, France) since October 2020 [16–18]. The ComCor survey aims to identify COVID-19 risk factors in the general population through a case-control study on community and occupational exposures to SARS-CoV-2.

Participants were included between 10 April and 9 July 2021. All laboratory-confirmed cases of COVID-19 (either nasopharyngeal reverse transcriptase–polymerase chain reaction (RT-PCR) or antigenic test) compiled by the French National Health Insurance were invited by email to complete a questionnaire following their positive test result. Respondents who selected the “healthcare worker or working within health field” criterion in the questionnaire were included as cases in this study. Controls were recruited during the same period through two different sources: 1) IPSOS, a French marketing research and public opinion specialist, selected controls from a panel representative of the French population using frequency-matching with cases for age, sex, region, population density and week of inclusion for the Comcor survey; and 2) 24 professional corporations, scientific associations and medical platforms were asked to forward the questionnaire to their members. Participants declaring to be HCWs using the above-described criterion and reporting no previous symptoms or positive test were enrolled as controls. The final study population was obtained by randomly selecting cases and controls with a 1:1 ratio by exact case-control matching for 10-year age-category distribution, sex and residential region.

### Data Collection

Participants received online information about the study and gave consent for participation by completing the self-administered questionnaire. They opted-in without any incentives or reminders. Questionnaires covered the 10 days preceding symptom onset for cases (or testing if asymptomatic) and the 10 days preceding inclusion for controls. As previously described, the questionnaire covered sociodemographic characteristics (age, sex, residential region, household composition), health condition (prior medical history, COVID-19-immunization status) [16]. Occupational activities were assessed for cases and controls: professional category, size and type of healthcare setting, frequency of contacts with general patients and COVID-19 patients, contacts with colleagues at work, and PPE use for COVID-19 patient care during the previous 10 days. HCW professions were grouped in 4 categories: medical staff (physicians, residents, dentists, pharmacists, biologists), nurses, nurse’s assistants and other professions (including, among others, laboratory or imaging technicians, administrative staff, speech or physical therapists, social workers and opticians). To account for previous immunization against SARS-CoV-2, we classified participants as either “not immunized”, “partially immunized” or “fully immunized” [17, 19]. Participants without any documented previous COVID-19, and either not vaccinated or first-dose vaccinated within the 21 days preceding inclusion were considered “not immunized”. Participants included 14 days to 6 months after laboratory-confirmed COVID-19 infection or >7 days after a second vaccine dose (28 days for 1-dose regimen) were classified as fully immunized. Other participants were considered partially immunized.

### Statistical Analyses

Categorical variables are described by number (percentage). All statistical analysis were computed with R Studio v4.1.0 (R Foundation for Statistical Computing, Vienna, Austria). Cases and controls were matched with the Matching package. Univariable and multivariable conditional logistic regression to account for the matching strategy, adjusted to the week of inclusion, assessed relationships between variables and the outcome (COVID-19). Missing data were managed with multiple imputations by chained equations using the MICE package. To evaluate imputation effects on our results, supplementary analysis was done on a sample of fully completed questionnaires only, excluding individuals with missing data. To compare risk factors within healthcare-setting categories (hospitals, LTCFs and primary care), subgroup analyses were conducted on three population samples using the same 1:1 matching strategy for age, sex and residential region. Strengthening of Reporting of Observational Studies in Epidemiology (STROBE) reporting guidelines were followed.

### Ethical Considerations

The ComCor study was approved by the *Comité de Protection des Personnes* (CPP) *Sud Ouest et Outre Mer-1* on 21 September 2020. The data protection authority *Commission Nationale de l’Informatique et des Libertés* (CNIL) authorized data processing on 21 October 2020. CPP and CNIL accorded authorizations for substantial modification to recruit controls through professional societies and associations on 31 March 2021. Informed consent was obtained from all participants. The study is registered with ClinicalTrials.gov under the identifier NCT04607941.

## RESULTS

### Participants

Among 562 841 individuals with confirmed COVID-19 contacted by the French National Health Insurance (10 April–9 July 2021), 6% completed the questionnaire, including 3510 HCWs, and 1:1 matching paired 2076 cases to 2076 controls for analysis. Overall, data were missing for 126 (6%) cases and 25 (1%) controls. The weekly number of inclusions and confirmed COVID-19 cases reported in France throughout the study period are reported in Figure 2.

**Figure 1.**
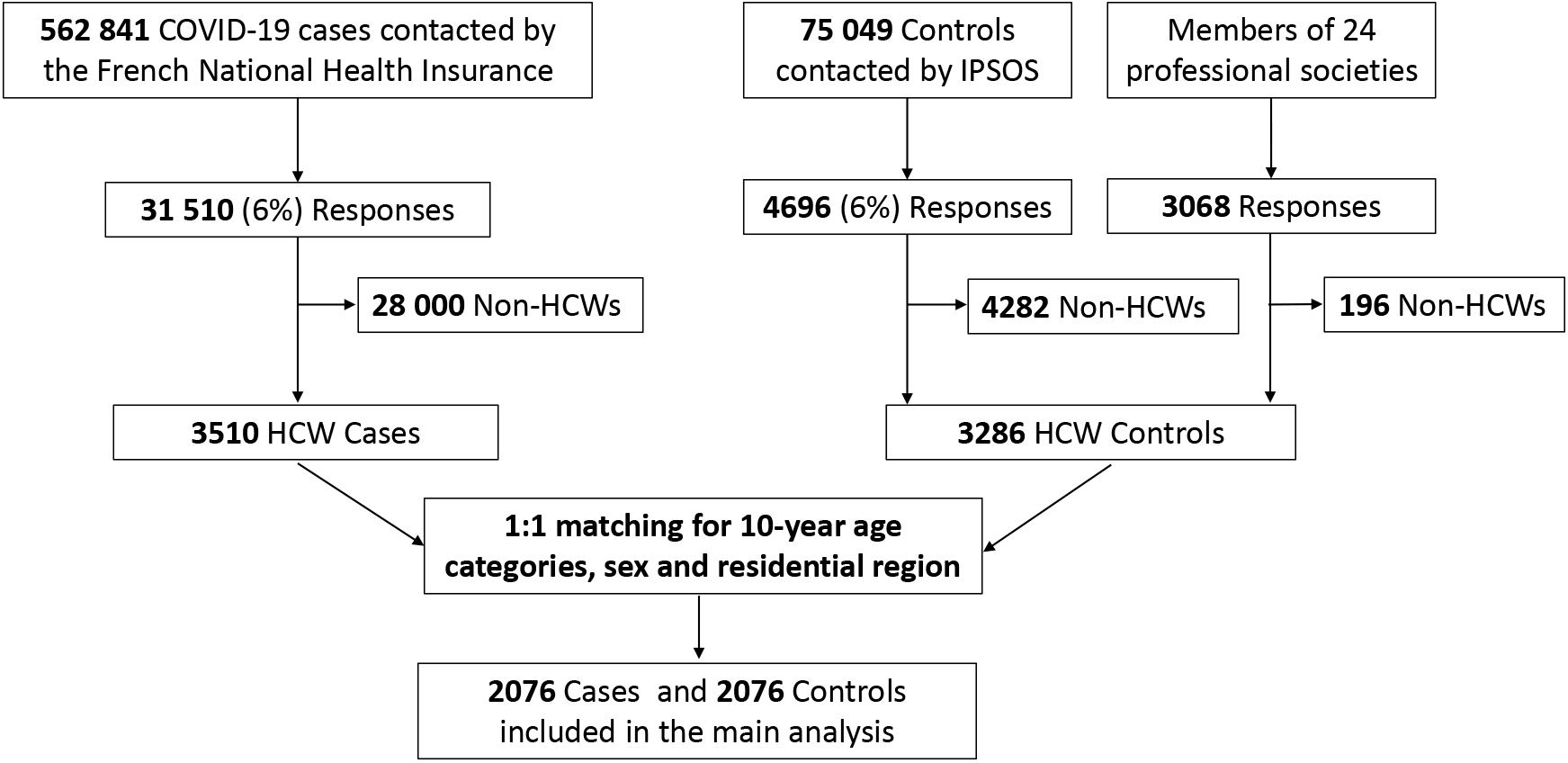
Flow chart of study participants. HCWs, healthcare workers; IPSOS, French marketing-research and public-opinion specialist.

**Figure 2.**
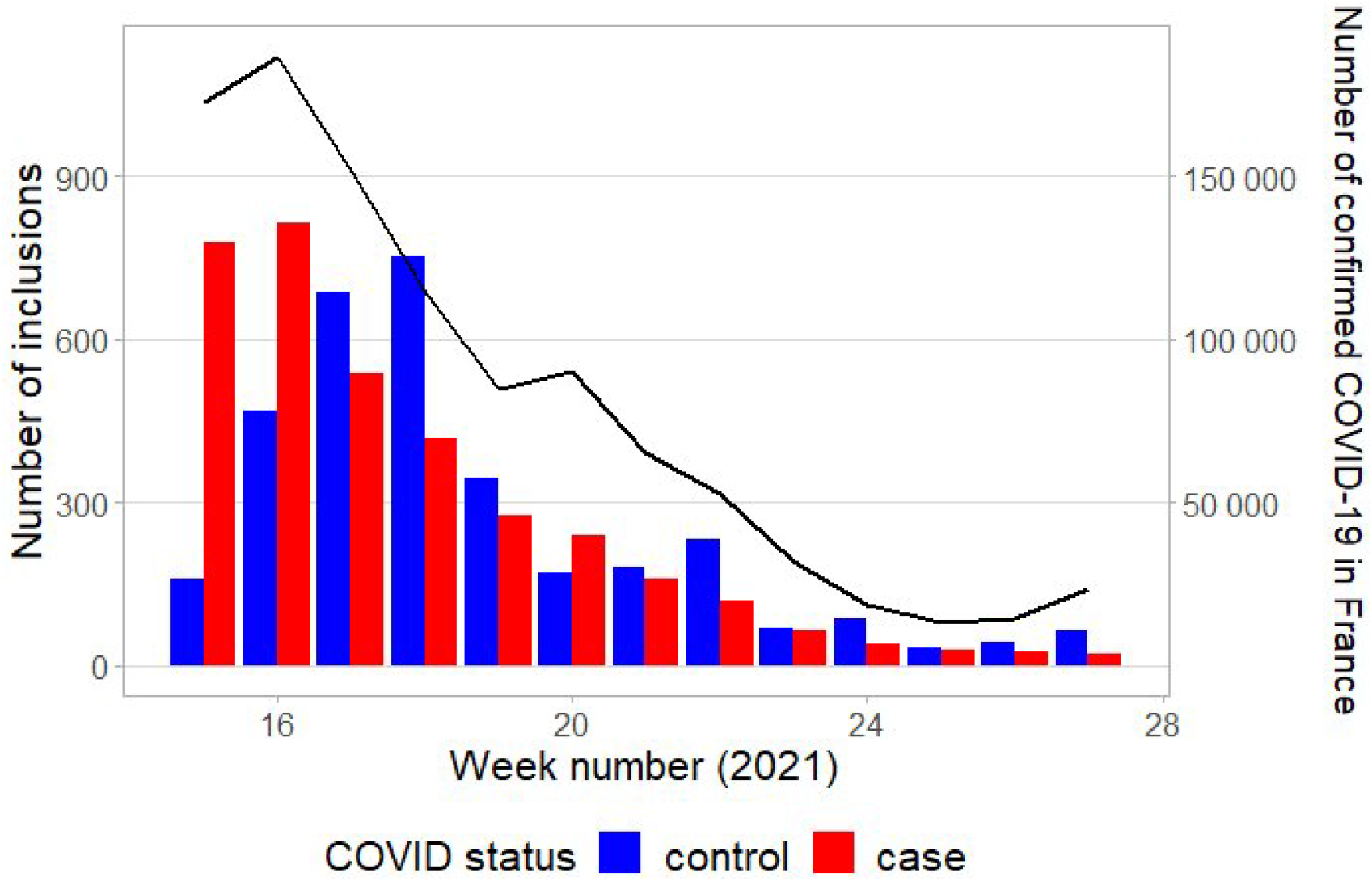
Weekly number of inclusions of cases (red bars) and controls (blue bars). The black line shows the weekly number of laboratory-confirmed cases reported in France throughout the study period (source: *Santé Publique France*).

Table 1 reports study population characteristics. Most participants were female, mostly working in primary care. Overall, 1770/4152 (43%) HCWs were classified as either partially or fully immunized against COVID-19; 678/4152 (16%) declared being posted in a COVID-19-dedicated unit or mostly caring for COVID-19 patients. In the subgroup of HCWs in contact with COVID-19 patients during the preceding 10 days (n = 2086), 1616 (77%) declared systematically wearing a gown, 1608 (77%) gloves, 1490 (71%) a N95 respirator, 1345 (64%) goggles/faceshield and 1146 (55%) an apron for patient care.

**Table 1.**
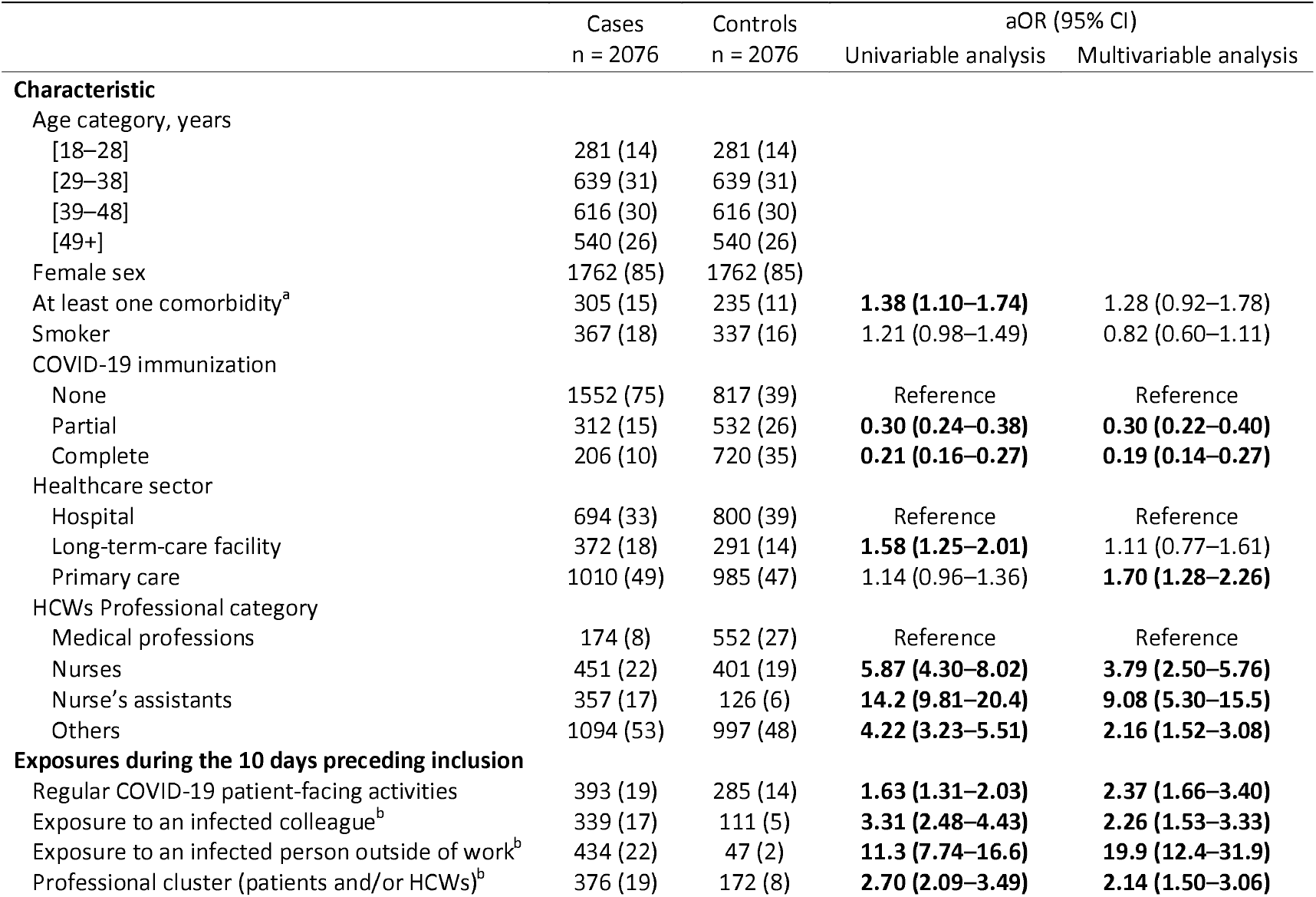
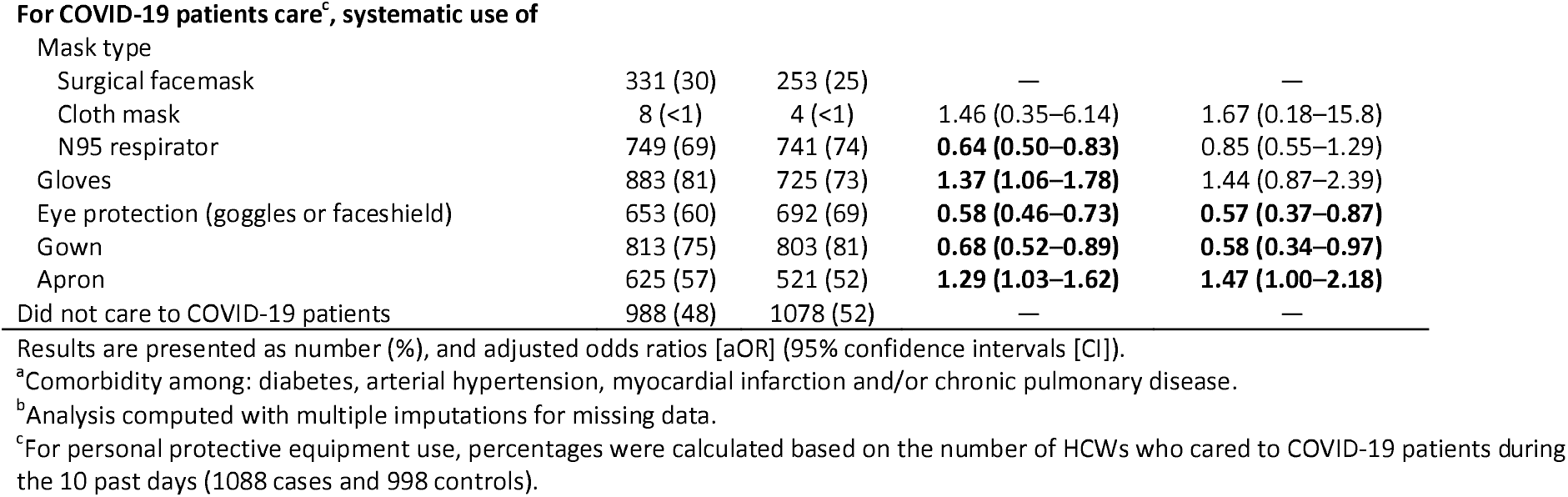
Study population and infection determinants: description and results of the univariable and multivariable conditional logistic regression analyses adjusted to the week of inclusion.

| Characteristic | Cases<br>n = 2076 | Controls<br>n = 2076 | aOR (95% CI) |  |
| --- | --- | --- | --- | --- |
|  |  |  | Univariable analysis | Multivariable analysis |
| **Characteristic** |  |  |  |  |
| Age category, years |  |  |  |  |
| [18–28] | 281 (14) | 281 (14) |  |  |
| [29–38] | 639 (31) | 639 (31) |  |  |
| [39–48] | 616 (30) | 616 (30) |  |  |
| [49+] | 540 (26) | 540 (26) |  |  |
| Female sex | 1762 (85) | 1762 (85) |  |  |
| At least one comorbidity<sup>a</sup> | 305 (15) | 235 (11) | **1.38 (1.10–1.74)** | 1.28 (0.92–1.78) |
| Smoker | 367 (18) | 337 (16) | 1.21 (0.98–1.49) | 0.82 (0.60–1.11) |
| COVID-19 immunization |  |  |  |  |
| None | 1552 (75) | 817 (39) | Reference | Reference |
| Partial | 312 (15) | 532 (26) | **0.30 (0.24–0.38)** | **0.30 (0.22–0.40)** |
| Complete | 206 (10) | 720 (35) | **0.21 (0.16–0.27)** | **0.19 (0.14–0.27)** |
| Healthcare sector |  |  |  |  |
| Hospital | 694 (33) | 800 (39) | Reference | Reference |
| Long-term-care facility | 372 (18) | 291 (14) | **1.58 (1.25–2.01)** | 1.11 (0.77–1.61) |
| Primary care | 1010 (49) | 985 (47) | 1.14 (0.96–1.36) | **1.70 (1.28–2.26)** |
| HCWs Professional category |  |  |  |  |
| Medical professions | 174 (8) | 552 (27) | Reference | Reference |
| Nurses | 451 (22) | 401 (19) | **5.87 (4.30–8.02)** | **3.79 (2.50–5.76)** |
| Nurse’s assistants | 357 (17) | 126 (6) | **14.2 (9.81–20.4)** | **9.08 (5.30–15.5)** |
| Others | 1094 (53) | 997 (48) | **4.22 (3.23–5.51)** | **2.16 (1.52–3.08)** |
| **Exposures during the 10 days preceding inclusion** |  |  |  |  |
| Regular COVID-19 patient-facing activities | 393 (19) | 285 (14) | **1.63 (1.31–2.03)** | **2.37 (1.66–3.40)** |
| Exposure to an infected colleague<sup>b</sup> | 339 (17) | 111 (5) | **3.31 (2.48–4.43)** | **2.26 (1.53–3.33)** |
| Exposure to an infected person outside of work<sup>b</sup> | 434 (22) | 47 (2) | **11.3 (7.74–16.6)** | **19.9 (12.4–31.9)** |
| Professional cluster (patients and/or HCWs)<sup>b</sup> | 376 (19) | 172 (8) | **2.70 (2.09–3.49)** | **2.14 (1.50–3.06)** |
| **For COVID-19 patients care<sup>c</sup>, systematic use of** |  |  |  |  |
| Mask type |  |  |  |  |
| Surgical facemask | 331 (30) | 253 (25) | — | — |
| Cloth mask | 8 (<1) | 4 (<1) | 1.46 (0.35–6.14) | 1.67 (0.18–15.8) |
| N95 respirator | 749 (69) | 741 (74) | **0.64 (0.50–0.83)** | 0.85 (0.55–1.29) |
| Gloves | 883 (81) | 725 (73) | **1.37 (1.06–1.78)** | 1.44 (0.87–2.39) |
| Eye protection (goggles or faceshield) | 653 (60) | 692 (69) | **0.58 (0.46–0.73)** | **0.57 (0.37–0.87)** |
| Gown | 813 (75) | 803 (81) | **0.68 (0.52–0.89)** | **0.58 (0.34–0.97)** |
| Apron | 625 (57) | 521 (52) | **1.29 (1.03–1.62)** | **1.47 (1.00–2.18)** |
| Did not care to COVID-19 patients | 988 (48) | 1078 (52) | — | — |

Results are presented as number (%), and adjusted odds ratios [aOR] (95% confidence intervals [CI]).
<sup>a</sup>Comorbidity among: diabetes, arterial hypertension, myocardial infarction and/or chronic pulmonary disease.
<sup>b</sup>Analysis computed with multiple imputations for missing data.
<sup>c</sup>For personal protective equipment use, percentages were calculated based on the number of HCWs who cared to COVID-19 patients during the 10 past days (1088 cases and 998 controls).

### Association between Exposures and COVID-19 Status

According to the multivariable analysis, the strongest predictor of contracting COVID-19 was exposure to an infected person outside work, while complete or partial immunization was protective (Table 1). Occupational exposure to an infected colleague, to COVID-19 patients, or working in a unit harboring a cluster of nosocomial cases increased the risk of HCW infection. Compared to medical staff, being a nurse or a nurse’s aide was significantly associated with the risk of contracting COVID-19. Eye protection (goggles or faceshield) and gowning for COVID-19-patient care were associated with lower risk, while wearing an apron posed a higher risk. N95-respirator-conferred protection was comparable to that of surgical facemasks. The supplementary analysis of cases with fully completed questionnaires (Supplementary Table 1) yielded similar results.

### Subgroup and Sensitivity Analyses

The subgroup analyses by healthcare sector are reported in Supplementary Tables 2 and 3. After 1:1 matching, 1388 HCWs from hospitals, 558 from LTCFs and 1842 from primary care were included. When caring for COVID-19 patients, HCWs declared more frequently wearing N95 respirators in hospitals and primary care than in LTCFs. Adherence to eye protection was particulary poor in primary care (46% of cases and 53% of controls). According to the multivariable analysis, partial or complete immunization was protective in all three settings, while exposure to an infected person outside work was again the main risk factor for infection.

## DISCUSSION

In this large case-control study, the strongest predictor of HCW COVID-19 infection was exposure to an infected person outside work. Contact with an infected colleague and regular COVID-19 patient-facing activities were also significantly, but to a lesser extent, associated with infection. Eye protection and gowning during patient care decreased the risk, while N95 respirators did not confer better protection than surgical masks. These results were consistent across healthcare settings (hospital, LTCFs and primary care).

As also shown by others, our results suggest that direct contact with infected household members, relatives or, to a lesser degree, colleagues were the main sources of HCW acquisition of COVID-19 [5, 6, 9]. Nevertheless, COVID-19 patient-facing activities seem to further enhance the risk, although previous results were heterogenous [2, 5, 7, 9, 20, 21]. One explanation for this heterogeneity across settings and wards may be the various degrees of HCW education and training to follow IPC protocols and best practices. Correct PPE use by HCWs is essential to avoid contaminations during patient care. Since the start of the pandemic, French guidelines have recommended universal masking with surgical facemasks for general patient care, and N95-respirator use for aerosol-generating procedures [22]. For confirmed COVID-19 patients, additional PPE, such as eye protection, and gowns or aprons must be worn. Gloves are restricted to activities carrying a risk of exposure to body fluids.

Our findings highlighted marked divergence of PPE use from French guidelines, since the large majority of HCWs declared systematically wearing a N95 respirator and gloves when caring for COVID-19 patients. However, our finding that N95 respirators were not superior to surgical facemasks for protecting HCWs during standard care is consistent with the results of a recent meta-analysis of 4 randomized controlled trials on other viral respiratory infections [11]. In a multicenter observational study in Switzerland, an institutional policy of systematic N95-respirator use was not associated with a lower HCWs’ seroconversion rate for COVID-19 [23]. Although additional safety conferred by eye protection was also suggested in a recent meta-analysis [24], most clinical workers find faceshields and goggles uncomfortable, impair vision and interfere with work, probably contributing to poor adherence [25]. Unexpectedly, apron use was associated with a heightened risk of contamination, while gowns were protective. The pandemic led to a widespread use or reuse of homemade aprons during COVID-19-patient care, owed to gown shortage for which donning and doffing might be easier. These observed misuses of aprons and possible lack of personal protection suits may have led to an increased risk of cross contamination during care.

HCWs from different healthcare settings have participated in the COVID-19 response. In France, 72% of nursing homes had at least one COVID-19-infected resident in 2020 [26], and numerous devastating outbreaks were described worldwide [27]. Herein, HCWs from LTCFs and primary care tended to be at higher risk of infection, which probably reflects a lack of extensive training (associated with large-scale staff turnover), and limited access to PPE and diagnostic tests, with a globally lower awareness on the infectious risk and prevention measures. Support from hospitals and regional health authorities should be encouraged to continue staff training and ensure PPE supply. The risk of contracting COVID-19 was also influenced by the professional category: being nurses and nurse’s assistants was more closely associated with COVID-19 than medical staff. Although those associations might be biased by unbalanced case and control populations for professions, they might also reflect that nurses and nurse’s assistants were engaged in more prolonged and closer patient care than other professions. This higher risk of infection was described for domestic cleaners and porters, but not for nurses and nurse’s assistants to our knowledge [5, 10, 21]. The result associated with the “other profession” category must be interpreted with caution because of the heterogeneity of professions, but numbers were too small to fit a statistical model to each individual profession.

The strengths of this study are the large sample size, enabling exact matching of cases and controls for age, sex and residential region, adjustment to the week of inclusion, and the nationwide distribution of study participants. Notably, sources of infection according to healthcare-facility category has not previously been assessed. The main limitations of the study are the low response rate of cases and controls and the use of an online questionnaire, which may have resulted in selection biases towards younger participants, more comfortable with internet and French language. Second, the underrepresentation of some professional categories impaired subgroup analyses, despite specific occupational exposures, e.g., physiotherapists or speech therapists. Third, the data used were relied upon HCW declarations, potentially influenced by social desirability or memorization bias. Fourth, we did not rule out past or current asymptomatic infections among controls [28]. Nevertheless, our population was composed of HCWs, more likely to recognize COVID-19 symptoms and with 3–5-fold higher access to tests than the general population, then lowering the risk of classification bias [1]. Finally, the study took place between April and July 2021, during the third COVID-19 wave in France. HCWs might have been better prepared and protected than during the first wave, especially regarding PPE, and the Delta and Omicron variants emerged in France after the end of the study period. Omicron transmissibility is much higher than previous variants, which may affect the relative weights of transmission sources and appropriate PPE [29].

In conclusion, our study results indicated that, for HCWs, COVID-19 patient-facing activities increased the risk of getting infected, while colleague-related and mostly community exposures appear to represent higher risks. Moreover, they suggest that, when caring for COVID-19 patients, HCWs should wear a surgical facemask (apart from aerosols-generating procedures), eye protection and a gown. The protection conferred by gloving should be further explored.

## Data Availability

All data produced in the present study are available upon reasonable request to the authors

## Supplementary Data

Supplementary materials are available at Clinical Microbiology and Infections online.

