## Supplementary Table 1 ; Supplementary Table 2 ; Supplementary Table 3 for "SARS-CoV-2 Exposures of Healthcare Workers from Primary Care, Long-Term Care Facilities and Hospitals: A Nationwide Matched Case-Control Study"

**Supplementary Table 1.** Complete cases population and infection determinants: description and results of the univariable and multivariable conditional logistic regression analyses adjusted to the week of inclusion.

| | Cases n = 2001 | Controls n = 2001 | aOR (95% CI) Univariable analysis | aOR (95% CI) Multivariable analysis |
|---|---|---|---|---|
| **Characteristics** | | | | |
| Age categories, years | | | | |
| [18–28] | 269 (13) | 269 (13) | | |
| [29–38] | 618 (31) | 618 (31) | | |
| [39–48] | 595 (30) | 595 (30) | | |
| [49+] | 519 (26) | 519 (26) | | |
| Female sex | 1708 (85) | 1708 (85) | | |
| At least one comorbidity[a] | 301 (15) | 223 (11) | **1.47 (1.16–1.87)** | 1.26 (0.89–1.78) |
| Smoker | 355 (18) | 334 (17) | 1.04 (0.85–1.28) | **0.68 (0.50–0.91)** |
| COVID-19 immunization | | | | |
| None | 1471 (74) | 761 (38) | Reference | Reference |
| Partial | 316 (16) | 520 (26) | **0.33 (0.26–0.41)** | **0.32 (0.24–0.44)** |
| Complete | 206 (10) | 712 (36) | **0.21 (0.16–0.26)** | **0.16 (0.11–0.23)** |
| Healthcare sector | | | | |
| Hospital | 711 (36) | 802 (40) | — | — |
| Long-term care facility | 379 (19) | 261 (13) | **1.78 (1.39–2.29)** | 1.41 (0.97–2.05) |
| Primary care | 911 (46) | 938 (47) | 0.99 (0.83–1.18) | **1.55 (1.15–2.08)** |
| HCW Professional category | | | | |
| Medical professions | 171 (8) | 559 (28) | — | — |
| Nurses | 454 (23) | 386 (19) | **5.77 (4.25–7.83)** | **5.16 (3.37–7.89)** |
| Nurse’s aides | 364 (18) | 115 (6) | **15.2 (10.4–22.2)** | **9.32 (5.38–16.1)** |
| Others | 1012 (51) | 941 (47) | **4.00 (3.08–5.19)** | **2.43 (1.70–3.48)** |
| **Exposures during the 10 days preceding inclusion** | | | | |
| Regular COVID-19 patient-facing activities | 407 (20) | 288 (14) | **1.64 (1.32–2.04)** | **1.93 (1.34–2.79)** |
| Exposure to an infected colleague | 351 (18) | 108 (5) | **3.59 (2.70–4.79)** | **2.92 (1.95–4.38)** |
| Exposure to an infected person outside of work | 444 (22) | 44 (2) | **15.5 (10.1–23.8)** | **26.8 (16.0–44.7)** |
| Professional cluster (patients and/or HCWs) | 391 (20) | 177 (9) | **2.53 (1.98–3.23)** | **1.94 (1.36–2.77)** |
| **For COVID-19 patients[b] care, systematic use of** | | | | |

| | | | | |
|---|---|---|---|---|
| Mask type | | | | |
| Surgical facemask | 340 (30) | 255 (26) | — | — |
| Cloth mask | 8 (1) | 2 (<1) | 1.89 (0.34–10.7) | 1.26 (0.12–12.8) |
| N95 respirator | 770 (69) | 742 (74) | **0.65 (0.51–0.83)** | 0.84 (0.54–1.31) |
| Gloves | 905 (81) | 717 (72) | **1.50 (1.16–1.94)** | 1.32 (0.79–2.20) |
| Eye protection (goggles or faceshield) | 668 (60) | 691 (69) | **0.54 (0.43–0.68)** | **0.47 (0.31–0.73)** |
| Gown | 834 (75) | 800 (80) | **0.67 (0.51–0.87)** | 0.65 (0.38–1.11) |
| Apron | 642 (57) | 523 (52) | **1.43 (1.15–1.78)** | **1.62 (1.09–2.40)** |
| Did not care for COVID-19 patients | 883 (44) | 1002 (50) | — | — |

Results are expressed as n (%) and adjusted odds ratios (aOR) (95% confidence intervals (CI)).

[a]Comorbidity among: diabetes, arterial hypertension, myocardial infarction and/or chronic pulmonary disease.

[b]For personal protective equipment use, percentages were calculated on the number of HCWs who cared to COVID-19 patients in the ten past days (1118 cases and 999 controls). HCWs, healthcare worker ; PPE, personal protective equipment.

**Supplementary Table 2** Healthcare workers' use of personal protective equipment for COVID-19-patient care across the three healthcare settings during the 10 days preceding inclusion.

| | Hospital | | Long-term-care facility | | Primary care | |
|---|---|---|---|---|---|---|
| | Cases<br>n = 694 | Controls<br>n = 694 | Cases<br>n = 279 | Controls<br>n = 279 | Cases<br>n = 921 | Controls<br>n = 921 |
| Personal protective equipment | | | | | | |
| Mask | | | | | | |
| Surgical facemask | 83 (18) | 102 (22) | 67 (44) | 64 (43) | 151 (40) | 78 (25) |
| N95 respirator | 383 (82) | 355 (78) | 85 (56) | 84 (57) | 226 (60) | 237 (75) |
| Gloves | 409 (88) | 351 (77) | 127 (84) | 113 (76) | 258 (68) | 205 (65) |
| Eye protection (goggles or faceshield) | 346 (74) | 351 (77) | 86 (57) | 113 (76) | 173 (46) | 167 (53) |
| Gown | 408 (88) | 388 (85) | 102 (67) | 118 (80) | 228 (60) | 214 (68) |
| Apron | 324 (70) | 277 (61) | 92 (61) | 110 (74) | 138 (37) | 101 (32) |
| Did not care for COVID-19 patients | 228 (33) | 238 (34) | 127 (45) | 131 (47) | 544 (59) | 606 (66) |

Results are expressed as number (%).

**Supplementary Table 3.** Infection determinants of subgroup multivariable conditional logistic regression analysis according to healthcare setting.

| | Hospital<br>n = 694 cases/694 controls | Long-term–care facility<br>n = 279 cases/279 controls | Primary care<br>n = 921 cases/921 controls |
|---|---|---|---|
| **Characteristic** | | | |
| At least one comorbidity[a] | 1.76 (0.99–3.12) | 1.26 (0.40–3.96) | 0.90 (0.50–1.62) |
| Smoker | 0.77 (0.46–1.28) | 0.60 (0.24–1.50) | 1.04 (0.63–1.71) |
| COVID-19 immunization | | | |
| None | Reference | Reference | Reference |
| Partial | **0.15 (0.08–0.27)** | **0.09 (0.02–0.37)** | **0.34 (0.22–0.53)** |
| Complete | **0.12 (0.06–0.22)** | **0.13 (0.04–0.50)** | **0.16 (0.09–0.30)** |
| **Exposures during the 10 days preceding inclusion** | | | |
| Regular COVID-19-patient-facing activities | **1.95 (1.15–3.28)** | **0.19 (0.04–0.97)** | **3.64 (1.59–8.34)** |
| Exposure to an infected colleague | 1.18 (0.66–2.09) | 4.34 (0.97–19.4) | **6.94 (2.45–19.6)** |
| Exposure to an infected person outside of work | **15.7 (7.38–33.5)** | **18.4 (4.53–74.3)** | **19.8 (9.51–41.3)** |
| Professional cluster (patients and/or HCWs) | **2.08 (1.19–3.63)** | 3.85 (0.79–18.7) | 1.97 (0.97–3.99) |
| **For COVID-19-patients care, systematic use of** | | | |
| Mask type | | | |
| Surgical facemask | Reference | Reference | Reference |
| N95 respirator | 1.68 (0.79–3.56) | 1.23 (0.30–5.01) | 0.61 (0.28–1.32) |
| Gloves | 1.29 (0.58–2.87) | 0.97 (0.11–8.69) | 2.50 (0.97–6.45) |
| Eye protection (goggles or faceshield) | 0.90 (0.46–1.77) | 0.30 (0.04–2.12) | 1.03 (0.50–2.11) |
| Gown | 0.70 (0.30–1.62) | 0.33 (0.03–3.37) | 0.61 (0.27–1.41) |
| Apron | 1.18 (0.66–2.11) | 1.28 (0.25–6.47) | 1.58 (0.83–3.01) |

Results are presented as n (%), and adjusted Odds Ratios [aOR] (95% Confidence Intervals [CI]).

[a] Comorbidity among: diabetes, arterial hypertension, myocardial infarction and/or chronic pulmonary disease. HCWs, healthcare worker ; PPE, personal protective equipment.
